# Depressive symptoms are associated with cigarette smoking among tuberculosis patients in Botswana

**DOI:** 10.1101/2021.09.14.21263357

**Authors:** Alexandria Jones-Patten, Qiao Wang, Keneilwe Molebatsi, Thomas E. Novotny, Kamran Siddiqi, Chawangwa Modongo, Nicola M. Zetola, Bontle Mbongwe, Sanghyuk S. Shin

## Abstract

**Background:** Researchers have increasingly recognized the adverse effects of smoking on tuberculosis (TB) outcomes. Smoking may be a maladaptive coping mechanism for depression and anxiety among TB patients; however, this association has not yet been investigated.

**Design/Methods:** We conducted a cross-sectional study among newly diagnosed TB patients between January and December 2019 in Gaborone, Botswana, and evaluated factors associated with cigarette smoking. Using the Patient Health Questionnaire-9 and the Zung Self-Rating Anxiety scale, we collected depression and anxiety scores, respectively; scores of ≥10 indicate depression and scores of ≥36 indicate anxiety. We performed Poisson regression analyses with robust variance to examine whether depression and anxiety were associated with smoking.

**Results:** One hundred and eighty participants with TB were enrolled from primary health clinics. Among all enrollees, depression was reported in 46 (27.1%) participants, while anxiety was reported in 60 (44.4%) participants. Overall, 45 (25.0%) participants reported current smoking, and the median number of cigarettes per day was 10. Depressive symptoms were associated with a higher prevalence of smoking (aPR: 1.82; 95% CI = 1.11, 3.01) after adjusting for sex, HIV status, food insecurity, anxiety, and income. The association between anxiety symptoms and cigarette smoking did not reach statistical significance (aPR 1.26; 95% CI: 0.78-2.05).

**Conclusions:** We found the association between depressive symptoms and smoking among TB patients in Botswana to be significant and the association between anxiety symptoms and cigarette smoking insignificant. Future studies should further investigate these associations when addressing TB care.

## Introduction

An estimated 1.4 million people died from tuberculosis (TB) globally in 2019; it is one of the top ten global causes of death worldwide [1]. Tobacco smoking is associated with an increased risk of TB infection and TB disease [2]. Research suggests smoking is associated with more extensive lung disease at time of TB diagnosis [3], higher rates of TB infection [4], and higher rates of TB treatment failures [5]. Despite evidence showing these harmful links between smoking and TB, the World Health Organization [6] estimates that 1.3 billion people use tobacco globally, and more than 80% of smokers live in low- and middle-income countries [7]. In Botswana, an upper middle-income country, 18% of adults use tobacco and 14.2% of adults smoke tobacco [8]. Further, depression is reportedly higher in TB patients than the general population [9]. Studies suggest an association between depression and smoking [10-12] as well as between anxiety and smoking [13, 14]; however, to our knowledge, these associations have not been studied in TB-infected populations. With both smoking and depression being more common in people with TB than in the general population [15], their association may be even stronger with potential adverse effects on patient outcomes. This study explores the association between smoking and common mental health issues among TB patients, including depression and anxiety.

## Methods

### Study Design, Population and Site Description

The methods for this study of TB patients have been described in detail elsewhere [16, 17]. Briefly, in an effort to explore the association between depression, anxiety and cigarette smoking in TB patients, this cross-sectional study was conducted from January to December, 2019 at 12 primary health care facilities in Gaborone, Botswana. Participants were eligible for inclusion if they were at least 18 years of age and older, with a microbiologically confirmed TB diagnosis, and were recruited by our research staff at the time of receiving TB diagnosis. All patients diagnosed with TB at the participating facilities were consecutively enrolled for this study. All TB patients regardless of disease site and drug susceptibility status were eligible.

### Measures

The primary outcome variable was smoking status. For analysis of smoking habits, participants were asked, “Are you a current smoker?” Current smoking was self-reported by participants, in which case participants were categorized into two groups: smokers and non-smokers. The independent variables were depression and anxiety. Depressive symptoms were assessed using the Patient Health Questionnaire-9. This nine-item instrument analyzes frequency of depressive symptoms up to two weeks prior to taking the questionnaire, using a 4-point Likert scale. Scores of 10 or higher indicate depressive symptoms [18]. The internal reliability of the Patient Health Questionnaire-9 in the current study was acceptable, with a Cronbach’s alpha of 0.74. This instrument has been validated in primary care clinic settings [18]. The Zung Self-Rating Anxiety Scale is a 20-item self-rating instrument which assesses cognitive, autonomic, motor and central nervous system symptoms of anxiety using a 4-point Likert-type scale ranging from ‘a little of the time,’ ‘some of the time,’ ‘good part of the time,’ and ‘most of the time’ [19]. Scores of 36 or higher on the Zung scale was indicative of anxiety symptoms. The Zung scale demonstrated acceptable internal reliability with a Cronbach’s alpha of 0.80 in this study.

We also administered a questionnaire to collect data on age, sex, income, education, employment status, history of alcohol use, and smoking status. Clinical information collected included TB diagnosis, TB symptoms and their duration, HIV status and antiretroviral treatment history, and CD4 counts (from the national HIV database for HIV-infected participants). Rifampin-resistant results from Xpert MTB/RIF were available for some participants.

Food insecurity was evaluated by the Household Food Insecurity Access Scale. This nine-item questionnaire distinguishes food-insecure households and determines the degree of food insecurity using a Likert-type scale that ranged from ‘never’, ‘rarely (1 or 2 times)’, ‘sometimes (3–10 times)’, and ‘often (more than 10 times)’.

### Data Collection

Potential participants were identified first from the lab and clinic register and were approached for recruitment by Research Assistants, who then screened them for eligibility and obtained informed consent. The Research Assistants read consent materials in English and/or Setswana (official languages in Botswana) to patients who were illiterate; all participants verified their consent to participate by signature. Data on socioeconomic, demographic characteristics and clinical information were collected in one-time interviews using the standardized questionnaires described above, prepared in both English and Setswana.

Participants whose total depressive symptom score corresponded with mild depression (score 5-9) were given a contact sheet for psychiatric services to seek help if symptoms worsened or persisted. Research Assistants helped make appointments for participants scoring a 10 or higher on the Patient Health Questionnaire-9 or those presenting with suicidal ideation. This study was approved by the Institutional Review Board of the University of California, Irvine and Botswana Ministry of Health Human Research Development Committee.

### Statistical Analysis

We investigated whether depression or anxiety was associated with the primary outcome of interest, smoking status, in people with TB. Participants were categorized into two exposure groups: current smokers and non-smokers. Participants were identified as experiencing depressive and anxiety symptoms if they scored ≥10 on Patient Health Questionnaire-9 and ≥36 on the Zung scale, respectively. To control for potential confounding, participants were categorized as experiencing ‘food security or mild insecurity’ or ‘moderate to severe insecurity [20]. We also dichotomized participants into having “no monthly income” and “some income,” and having “no education” and “some education,” as the variation between these categories was minimal. We examined the bivariate associations of current smoking with age, education, sex, monthly income, HIV status, food insecurity, anxiety, and depression. Poisson regression modeling with robust variance was used to estimate prevalence ratios (PRs) and 95% confidence intervals (95% CI) for bivariate and multivariable analysis with current smoking status specified as the outcome variable [21, 22]. The multivariate Poisson regression model assessed the association between mental health disorders and current smoking status adjusting for sex, income, HIV status, and food insecurity. A sensitivity analysis was also conducted; the analysis was restricted to men only as there were very few women identifying as smokers. We also conducted a sensitivity analysis to assess whether alcohol consumption would confound our results. Data were analyzed using SAS version 9.4 (SAS Institute, Cary, North Carolina, U.S.).

## Results

A total of 180 participants were included in the study; the majority were male (64.4%, n=116), and the mean age was 37 years. Among all enrollees, depression was reported in 46 (27.1%) participants, while anxiety was reported in 60 (44.4%) participants. One quarter (n=45) of participants identified as smokers. Smokers were more likely to be male (91.1%, n=41), and smoked a median of ten cigarettes per day (67%, n=30). Compared to non-smokers, current smokers were more likely to report depressive symptoms (PHQ-9 ≥10: 40.0% vs. 21.5%) and anxiety symptoms (Zung ≥ 36: 55.6% vs. 44.4%). Moderate to severe food insecurity was also reported more in smokers than non-smokers (48.9% vs. 34.8%), as was alcohol use (100% vs. 25.2%) respectively (Table 1).

**Table 1.** Characteristics of study participants in Gaborone, Botswana, 2019 (n=180)

| Characteristic |  | Non smoker<br>n/N (%) or median<br>(IQR) | Smoker<br>n/N (%) or median<br>(IQR) |
| --- | --- | --- | --- |
| Overall |  | 135/180 (75.0) | 45/180 (25.0) |
| Age (in years) |  | 36.0 (27.0-46.0) | 39.0 (30.0-43.0) |
| Number of cigarettes<br>smoked per day |  | NA | 10.0 (5.0-20.0) |
| HIV | Negative | 60/135 (44.4) | 21/45 (46.7) |
|  | Positive | 75/135 (55.6) | 24/45 (53.3) |
| Food insecurity | None or mildly<br>insecure | 88/135 (65.2) | 23/45 (51.1) |
|  | Moderate to<br>severe | 47/135 (34.8) | 22/45 (48.9) |
| Depression | No | 106/135 (78.5) | 27/45 (60.0) |
|  | Yes | 29/135 (21.5) | 18/45 (40.0) |
| Anxiety | No | 75/135 (55.6) | 20/45 (44.4) |
|  | Yes | 60/135 (44.4) | 25/45 (55.6) |
| Education | No | 12/135 (8.9) | 5/45 (11.1) |
|  | Yes | 123/135 (91.1) | 40/45 (88.9) |
| Income | No | 55/135 (40.7) | 10/45 (22.2) |
|  | Yes | 80/135 (59.3) | 35/45 (77.8) |
| Gender | Male | 75/135 (55.6) | 41/45 (91.1) |
|  | Female | 60/135 (44.4) | 4/45 (8.9) |
| Alcohol | No | 101/135 (74.8) | 0 |
|  | Yes | 34/135 (25.2) | 45/45 (100.0) |
| CD4* (cells/mm<sup>3</sup>) | ≤ 200 | 23/75 (30.7) | 7/24 (29.2) |
|  | > 200 | 40/75 (53.3) | 15/24 (62.5) |
|  | Missing | 12/75 (16.0) | 2/24 (8.3) |
| ART* | Never taken<br>ART | 25/75 (33.3) | 6/24 (25.0) |
|  | Taking ART | 45/75 (60.0) | 14/24 (58.3) |
|  | Took ART but<br>stopped | 5/75 (6.7) | 4/24 (16.7) |

Abbreviations: IQR=Interquartile range; ART=Antiretroviral therapy
\*Among HIV-positive patients only (n = 99)

In the unadjusted model, we found female participants were less likely than male participants to be current smokers (cPR: 0.18; 95% CI: 0.07-0.47). Having income (cPR: 1.98; 95% CI: 1.05-3.73) and depressive symptoms (cPR:1.89; 95% CI: 1.15-3.10) were more likely to be associated with current smoking (Table 2). After adjusting for HIV co-infection, income, anxiety symptoms, and food insecurity, depressive symptoms (aPR: 1.82; 95% CI: 1.11-3.01) and male sex (aPR: 0.18; 95% CI: 0.07-0.48) remained to be significantly associated with current smoking (Table 2). Depression (aPR: 1.09; 95% CI: 1.03-1.15) and income (aPR: 2.15; 95% CI: 1.02-4.53), were each positively associated with smoking intensity. Compared to men, women demonstrated lower smoking intensity (aPR: 0.18; 95% CI: 0.07-0.48). In our sensitivity analyses, we found similar results for male participants only, (aPR: 2.22, 95%CI: 1.41-3.48), and a reduced level of association among alcohol consumers only (aPR: 1.44, 95% CI: 1.00-2.07) for the association between depression and smoking. The association between anxiety symptoms and cigarette smoking did not reach statistical significance (aPR 1.26; 95% CI: 0.78-2.05).

**Table 2.** Correlates of current smoking (n=180)

| Characteristic |  | Crude PR<br>(95% CI) | Adjusted PR<br>(95% CI) |
| --- | --- | --- | --- |
| HIV | Negative | 1.00 | 1.00 |
|  | Positive | 0.94 (0.56-1.55) | 0.87 (0.54-1.39) |
| Food insecurity | None or mildly insecure | 1.00 | 1.00 |
|  | Moderate to severe | 1.54 (0.93-2.54) | 1.43 (0.85-2.40) |
| Depression | No | 1.00 | 1.00 |
|  | Yes | **1.89 (1.15-3.10)** | **1.82 (1.11-3.01)** |
| Anxiety | No | 1.00 | 1.00 |
|  | Yes | 1.40 (0.84-2.33) | 1.26 (0.78-2.05) |
| Education | No | 1.00 | Not included |
|  | Yes | 0.83 (0.38-1.83) |  |
| Income | No | 1.00 | 1.00 |
|  | Yes | **1.98 (1.05-3.73)** | 1.67 (0.92-3.04) |
| Gender | Male | 1.00 | 1.00 |
|  | Female | **0.18 (0.07-0.47)** | **0.18 (0.07-0.48)** |
| Age |  | 1.00 (0.99-1.02) | Not included |

Abbreviations: PR=Prevalence ratio; CI=Confidence interval

## Discussion

In this study, depression was associated with current smoking among TB patients after adjusting for potential confounders. Additionally, we found that men were more likely to engage in smoking than women in this study. Patients with TB have reported both depression [23, 24] and smoking during treatment of TB [3, 25], with both potentially leading to reduced treatment success. In addition to the hypothesis that this relationship could be bidirectional, another hypothesis for the association between depression and smoking is the desire to self-medicate [26]. Nicotine causes the release of dopamine; when smoking stops, depressed mood can occur as a result of nicotine withdrawal [27]. The negative effect of depression may be enhanced among people with TB, as treatment for TB can take a minimum of six to nine months. Our findings suggest that current efforts to help TB patients quit smoking may benefit from the management of multi-morbidity, including mental health and smoking cessation counseling.

The findings of this study should be considered with several limitations. First, these data are from a cross-sectional study, leaving the direction of causality between smoking and depression unclear. However, other studies show consistent associations between depression and smoking [28, 29]. Given the high proportion of smoking and depression among TB patients, a more consistent approach would be addressing the multi-morbidities of smoking and depression in TB treatment programs. Additionally, data about recent quit attempts was not collected. It is possible people recently diagnosed with TB could have quit smoking upon learning of their TB diagnosis and therefore classified as non-smokers. Further, because we used a convenience sample in Botswana for this study, our results may not be generalizable to other TB populations. Finally, some data collected in our study were based on self-report (anxiety scores, smoking status), which may lead to misclassification of variables included in our model. However, research staff were trained to minimize this bias via objective interview techniques; for questions related to mental health disorders, participants were able to directly record their answers on the questionnaire.

## Conclusion

This study expands our understanding of the association between depression and smoking status among TB patients in Botswana. Additionally, this study adds evidence to the need for specific public health efforts including smoking cessation interventions as part of TB treatment programs. Previous programs have evaluated smoking cessation as part of the core treatment program for people with TB [30], and should be considered in future TB treatment programs. Mental health screenings should also be included in treatment of TB to provide person-centered care among these higher risk patients and further develop treatment plans for patients with TB multimorbidity. Future studies should employ larger sample size in different settings to validate our findings. Additionally, future research should review the course of depression and anxiety and its association with smoking cessation during TB treatment [31].

## Statements

### Informed Consent

Informed consent was obtained from all individual participants included in the study.

### Ethical Approval

All procedures performed in studies involving human participants were in accordance with the ethical standards of the institutional and national research committee and with the 1964 Helsinki declaration and its later amendments or comparable ethical standards. This study was approved by the UCI IRB and the Botswana Ministry of Health Human Research Development Program.

### Welfare of Animals

This article does not contain any studies with animals performed by any of the authors.

## Data Availability

We have deposited the research dataset, which is available in Dryad.

## Acknowledgements

We would like to thank study participants for their time and efforts. We would also like to thank University of California, Irvine and Botswana Ministry of Health and Wellness Human Research Development Committee. Dr. Siddiqi was funded by Medical Research Council (UK) from TB Multimorbidity Grant (Ref No: MC_PC_MR/T037806/1).

## Conflict of Interest Declaration

All authors declare we have no conflict of interest.

## Notes

### Competing Interest Statement

The authors have declared no competing interest.

### Author Declarations

This study was approved by the Institutional Review Board of the University of California, Irvine and Botswana Ministry of Health Human Research Development Committee.

